# Synthesizing evidence on the impacts of COVID-19 regulatory changes on methadone treatment for opioid use disorder: Implications for U.S. federal policy

**DOI:** 10.1101/2022.12.15.22283533

**Authors:** Noa Krawczyk, Bianca D. Rivera, Emily Levin, Bridget C.E. Dooling

**Author notes:** **Corresponding Author** Noa Krawczyk, PhD, Assistant Professor, Center for Opioid Epidemiology and Policy (COEP), Department of Population Health, NYU Grossman School of Medicine.

## Abstract

As the U.S. faces a worsening overdose crisis, improving access to evidence-based treatment for opioid use disorder (OUD) remains a central policy priority. Federal regulatory changes in response to the COVID-19 pandemic significantly expanded flexibilities on take-home doses for methadone treatment for OUD. These changes have fueled critical questions about the impact of new regulations on OUD outcomes, and the potential health impact of permanently integrating these flexibilities into treatment policy going forward. To aide US policy makers as they consider implementing permanent methadone regulatory changes, we conducted a review synthesizing peer-reviewed research evidence on the impact of the COVID-19 methadone-take-home flexibilities on methadone program operations, OUD patient and provider experiences, and patient health outcomes. We interpret this evidence in the context of the federal rulemaking process and discuss avenues by which these important findings can be incorporated and implemented into U.S. substance use treatment policy going forward.

## 1. Introduction

One million lives have been lost to the overdose crisis that has ravaged U.S. communities for two decades.^1^ Exacerbated by the COVID-19 pandemic, 2021 was the deadliest year of this crisis to date, with over 100,000 deaths due to overdose.^2–4^ A central challenge of the overdose crisis, both before and during the pandemic, has been limited access to life-saving treatments with medications for opioid use disorder (MOUD), including methadone, buprenorphine, and extended-release naltrexone.^5^ These pharmacological treatments are highly effective at reducing overdose risk,^6^ and improving many other health and social outcomes.^7^ Of the three MOUD, methadone has the most extensive evidence base and has been used successfully for treatment of OUD since the 1960s.^8^ However, in 2020, only 311,000 people received methadone, less than 5% of the 7.6 million individuals estimated to have OUD.^9,10^

Lack of methadone utilization is largely attributed to the rigid and burdensome structure by which methadone treatment is regulated and delivered in the U.S. Heavily influenced by racialized Drug War rhetoric during the 1970s, the U.S. system only allows methadone to be delivered via specialty opioid treatment programs (OTPs) that are subject to stringent regulations by the Drug Enforcement Administration and Substance Abuse and Mental Health Services Administration (SAMHSA).^11,12^ Citing concern for abuse, diversion, and risk of overdose, these regulations limit the number of doses given to patients and prohibit entities like pharmacies from playing a role.^13^ The result is a system that requires patients to make almost-daily visits to an OTP to receive medication, except on days in which the clinic is closed when patients can take a dose home. Patients can qualify for additional “take-home” doses, but this can take months or years, and often depends on many subjective factors decided upon by clinic staff.^12^

This system is especially burdensome for patients who live far from OTPs, lack transportation, or have competing work or childcare responsibilities.^14^ Additionally, this system disproportionately impacts racially minoritized communities, where there is less access to office-based buprenorphine, a much less heavily regulated MOUD.^15^ Decades of research document experiences of patients who describe the OTP system, and the daily visits in particular, as burdensome, degrading and dehumanizing, often acting as a deterrent from initiating or staying in treatment.^14,16^

In response to the COVID-19 pandemic, federal regulators in the U.S. issued a suite of policy changes to support social distancing in healthcare.^17^ In mid-March 2020, SAMHSA issued guidance allowing states to request flexibility for OTPs to give additional take-home doses of methadone. Under this policy, “stable” patients could receive 28 days of medication and “less stable” patients could receive up to 14 days of medication.^18^ In 2021, SAMHSA announced plans to make the pandemic flexibilities permanent. Long-standing federal policy holds that as agencies prepare to issue new rules, they should draw upon the best reasonably obtainable scientific information to inform their policies (EO 12866).^19^ A common challenge regulators face in justifying proposed policy changes is not having data. In this case, SAMHSA can benefit from dozens of studies that explored the impacts of the pandemic flexibilities for methadone take-homes. In this Health Policy Review, we aim to: 1) Extract, review and synthesize published research evidence on the impact of the COVID-19 methadone-take-home flexibilities on OTP program operations, patient and provider experiences, and patient health outcomes; 2) interpret research findings in the context of the U.S. federal rulemaking process; and 3) discuss avenues by which findings can be incorporated and implemented into updated federal regulations.

## 2. Methods

### a. Search strategy

Our study team combines public health and regulatory expertise. For this review we searched for peer-reviewed studies published online or in-print between March 1, 2020 and September 6, 2022 focused on mesauring the effects of SAMHSA’s pandemic guideline. We searched PubMed, PsycInfo, and Google Scholar with combinations of the following terms: COVID-19, pandemic, methadone, take-home, methadone maintenance therapy/MMT, opioid treatment program/OTP, opioid-related disorder, and opiate substitution treatment (see Appendix Table 1 for search strategy). We also reviewed reference lists from included articles for relevant studies not identified by the database search.

**Table 1:** Characteristics of Included Studies.

|  | n (%) |
| --- | --- |
| **Study Design<sup>a</sup>** |  |
| Observational outcomes study | 12 (41·38) |
| Qualitative study | 11 (37·93) |
| Closed end survey | 4 (13·79) |
| Randomized trial | 1 (3·45) |
| Quantitative content analysis<sup>b</sup> | 2 (6·9) |
| **Setting** |  |
| Single OTP | 7 (24·14) |
| Multiple OTPs | 13 (44·83) |
| Other<sup>c</sup> | 9 (31·03) |
| **U.S. Region** |  |
| Northeast | 7 (24·14) |
| South | 3 (10·34) |
| Midwest | 1 (3·45) |
| West | 6 (20·69) |
| Multistate<sup>d</sup> | 12 (41·38) |
| **Research Question Addressed<sup>a</sup>** |  |
| Implementation | 17 (58·62) |
| Patient treatment experience and quality of life | 10 (34·48) |
| Provider experiences and attitudes towards care | 7 (24·14) |
| Overdose | 6 (20·69) |
| Illicit drug use & diversion | 8 (27·59) |
| Treatment initiation & retention | 1 (3·45) |

*Notes.*
OTP= Opioid Treatment Program
<sup>a</sup>Not mutually exclusive categories as studies may have multiple outcomes or mixed methods designs
<sup>b</sup>Quantitative content analyses used natural language processing and machine learning to evaluate outcomes
<sup>c</sup>Studies from settings other than OTPs include the following data sources: Reddit forums (4), convenience sample of PWUD, clinicians, and government officials (3), and the National Poison Data System (1).
<sup>d</sup>Multi-state studies include participants from more than one state, including the studies using Reddit and the National Poison Data System as data sources. While Reddit is an international platform, we used findings from studies that referred to the US methadone treatment system.

### b. Screening and data extraction

Articles were included if they were: (1) English-language and U.S.-based, (2) original research, (3) measuring the role or effect of the SAMHSA guidance. We excluded any articles focusing solely on pre-pandemic outcomes. Using Covidence, a subscription-based systematic review tool,^20^ we removed duplicates, screened titles and abstracts for relevance and then reviewed the full-text to assess eligibility based on inclusion criteria. The full study team conferred to select the final list of eligible articles. Study members then extracted findings on six research questions with policy relevance: how the new methadone take-home flexibilities (1) were implemented; (2) influenced perceptions and experiences of methadone patients and (3) methadone providers; (4) affected overdose risk, (5) illicit drug use and methadone non-compliance or diversion, and (6) affected methadone treatment initiation and retention.

### c. Synthesis of findings in the context of federal rulemaking

Our team first reviewed and synthesized findings related to each of the research questions, considering the different samples, study designs, analytic methods used, and limitations and strengths of each study. Our team then assessed the implications of the findings for the upcoming SAMHSA rulemaking. In the U.S., to write a new rule, federal regulators must generally follow certain steps set out in the Administrative Procedure Act: issue a proposed rule, take public comment, and then issue a final rule (5 U.S.C. 553). In a proposed rule, the regulator explains its proposed changes and also provides legal, policy, economic, and other justifications for the changes. The regulator’s task is not merely to describe the regulatory change, but also to explain why the change is consistent with the law and in the public interest. Our findings are therefore organized using the instructions for regulators outlined in Executive Order 12866,^19^ with a subsequent discussion of implications and implementation considerations.

## 3. Results

### a. Characteristics of reviewed studies

The search strategy resulted in 576 total articles, out of which 29 met full criteria for this review (Figure 1). Table 1 summarizes descriptive characteristics of the 29 articles, with detailed characteristics and outcomes by study available in Appendix Table 2. Most studies were qualitative (N=11, 38%) or observational outcome studies (N=12 (41%)), and took place across multiple OTPs (N=13, 46%) and multiple U.S. states (N=11, 39%). The most common outcome assessed was implementation of new take-home flexibilities (N=17, 59%), followed by methadone patient (N=10, 34%) and provider (N=7, 24%) experiences. To illustrate many of the patient and provider perspectives expressed, we include a subset of direct quotes extracted from qualitative studies, conveying some of the main emerging themes regarding experiences and perceptions (Table 2). Table 3 summarizes primary findings by each of our six policy-relevant questions.

**Figure 1:**
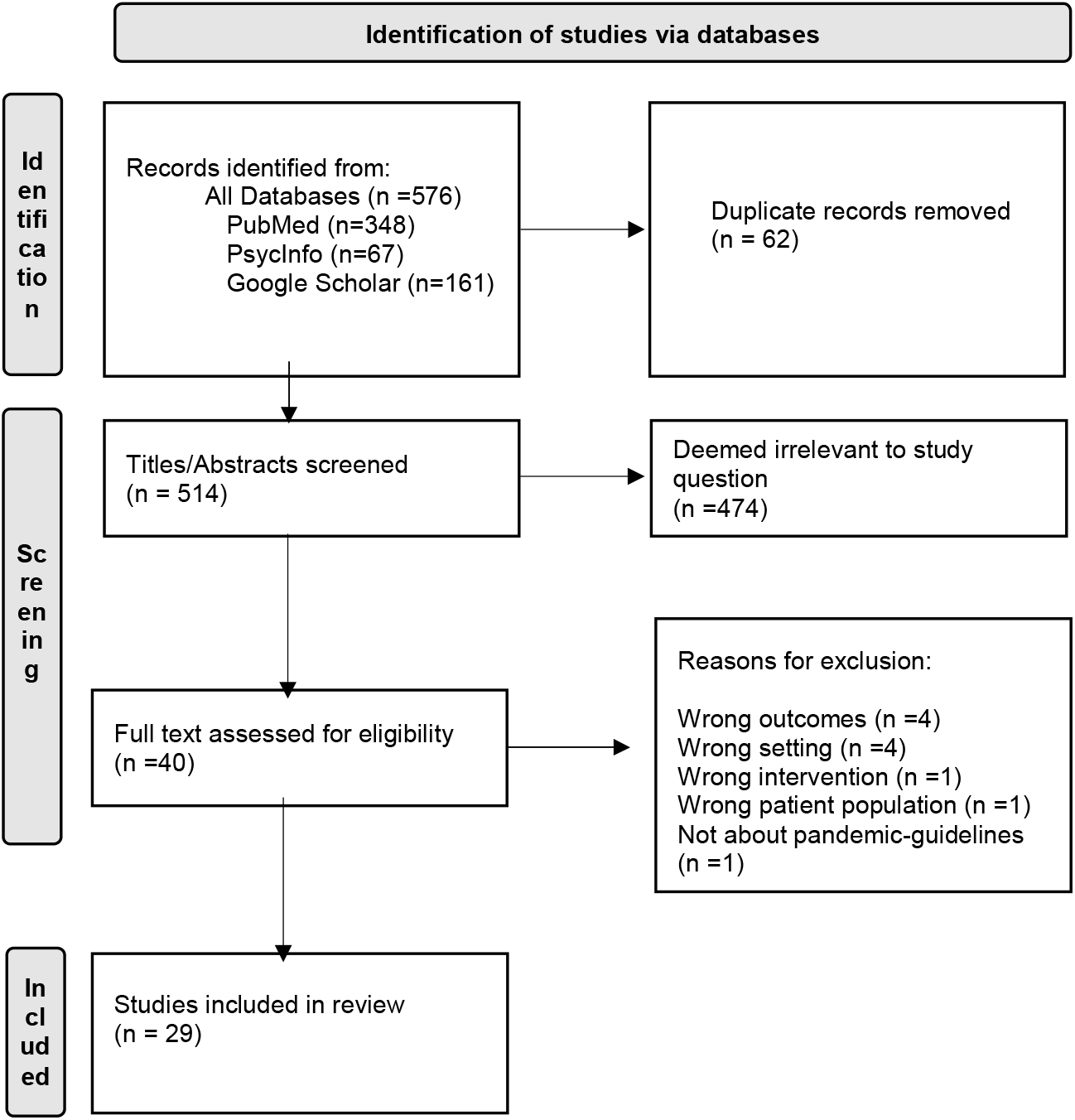
PRISMA^57^ diagram of studies considered and selected in the review.

**Table 2.** Select quotes from qualitative studies capturing patient and provider experiences, by theme:

|  |
| --- |
| **Theme 1: Patient challenges with implementation of pandemic flexibilities** |
| “I live with high risk people. I’m afraid that going to the clinic puts them at risk, but my clinic refuses to follow SAMHSA guidelines.” (Nobles, 2021) |
| “I still had to get up and go every day. They weren't running trains. They weren't running the buses...I'm five miles away from [the] inner city. And here I am having to fucking ride the bike down the highway...We couldn't do anything, but it's okay to send the drug addicts out. The homeless guys out so that they can go get their food stamps and fucking methadone” (Harris 2022) |
| “The most [take-homes] one can get at [my opioid treatment program] is 1 week. I asked if they would have to make special exceptions because of the [COVID-19] crisis. I was told that nobody has to do anything for us” (Sarker, 2022) |
| “My immune system is comprised. All documented via paperwork from doctors/emergency rooms plus my medication lists. I've not been given the take-home doses nor have others that have health issues causing us to be high risks [] for [COVID-19]. What can we do? I've talked to the drug board, and health department. As have others. We feel very afraid that we will die trying to get dosed. It's maddening knowing the clinic has permission to give take-homes but refuse. They just "blow us off" saying they will call etc[.] etc. But no one of us has been called yet.” (Sarker, 2022) |
| **Theme 2: Patient positive experiences with take-home flexibilities** |
| “...I feel that it's given me a sense of responsibility. I wasn't sure if I was ready to handle– but of course, I rose to the challenge. That makes me feel proud of myself. It really does. Having that responsibility and taking care of them on my own.” (Hoffman, 2022) |
| “[Take-homes make] it much easier for me– probably more than most people, because it might not be a big deal to a lot of people but I live twenty four miles... Every day when I was coming in, it was almost forty-five minutes to an hour driving round trip every day just the driving. I did that for probably a couple of years. Six days a week.” (Hoffman, 2022) |
| “I didn't feel nervous... that I would take them all at once or have trouble taking them every day. I didn't feel like I wasn't being monitored properly because I wasn't coming into the clinic all the time... When you get your take-homes it's like you feel you are being trusted to take care of yourself, and do the right thing...it felt great...that I was on the right track in my recovery.” – 39-year-old woman, P29 (Levander, 2021) |
| “[I am] able to live a normal life without having to come in every single day. I have a baby at home and stuff so that's initially why I joined the clinic, Not having to come in I feel a little more independent. I feel when I do get a job it will be a lot easier, just enjoy being able to be more like a normal person, just having my medication at home.” – 31-year-old woman (Levander, 2021) |
| “I was able to go camping with my mom and not have to worry about asking for extra doses. I went and saw my son and I didn't have to ask for extra doses 'cause I already had them. Just made it a little easier. A lot easier.” – 51-year-old woman (Levander, 2021) |
| “Before I would go every week. Now I'm going every two weeks...Definitely better...Well, the thing with work, not having to worry about being late for work on those days.” (Walters, 2022) |
| **Theme 3: Patient negative experiences with take-home flexibilities** |
| “Now, I like coming in everyday because I think it keeps you on track...I think it's better for people at first...I wasn't even getting take-homes, and all of a sudden here I am getting two weeks of my medicine so it was kind of a lot...For me it just wasn't good at the time because I was still pretty new in my sobriety, you have to trust in yourself and everybody is different.” – 44-year-old woman (Levander, 2021) |
| "I found [the methadone take-home] very hard to do because I would drink a little extra on day four, and it would leave me running on empty...So I basically told on myself and told [the clinic] that I was having trouble with the take-homes, so they stopped giving them to me...I like it better because [going to the clinic] gets me up and ready for the day. I get up early, so I'm not sleeping all day. So it gets me motivated" – 37-year-old man (Harris, 2022) |
| “When you're on the clinic, you go every single day, which means you got to get up and leave the house, and just go. Now, they were giving people take-homes, which means some people got three, and six, whatever. I ended up getting six bottles so I could stay home. In a way, it helped me, but then in a way it hurts too because I started that feeling again of not leaving the house...I think I probably shouldn't have got any take-homes and just continued going daily, and seeing the nurses and the counselors that were there.” - 52-year-old woman (Harris, 2022) |
| **Theme 4: Provider positive experiences with take-home flexibilities** |
| “Keeping it as loose as possible so that individual clinics could do what they think is clinically appropriate feels like it would be safer than the old ways of doing things. [ ... ] The idea [is] to help people achieve greater success and greater liberty from us. And it’s been okay. We haven’t had any terrible stuff from that.” (Suen, 2022) |
| “This was the most surprising thing... getting the take-home medications that they have not earned, actually motivated them to change that they are now meeting the criteria... So that for them it's no longer a pandemic bottle, it is another bottle that I have earned” (Trietler, 2022) |
| “But at the end of the day, a regulatory requirement that we must see people face-to-face for that visit, there is no question that there are people that will not be able to access care during that windowed timeframe. There will be people that overdose and die because, I mean, that will happen. We strongly advocate for that not being reinstated and that we are able to continue to deliver care” (Trietler, 2022) |
| “Our initial thinking that it was just going to be a complete mess... and it ended up not turning out that way at all” (Trietler, 2022) |
| “the levers for telemedicine, for take-home supplies of methadone have really been a game changer. And I’m really hoping that it’s something that is extended and we can move that up permanently” (Hunter, 2021) |
| **Theme 5: Provider negative experiences with take-home flexibilities** |
| “As a contingency management tool, we've lost the ability to grant or remove takehome dosages from patients, either as an incentive for doing better or as something they would lose if they did worse. So, we've definitely lost a lot of tools” (Trietler, 2022) |
| “A pregnant mother that I can recall, she just needed the contact that came with daily dosing. She needed the support that we were giving her, and the love and attention we were giving her that she wasn’t getting at home to help her get through her pregnancy... I like that accountability piece of coming in every day, so we have eyes on them. That’s why I prefer methadone over Suboxone.” (Madden, 2021) |

**Table 3.** Summary of findings by research question.

|  |  |
| --- | --- |
| **1. How did the new methadone take-home flexibilities become implemented in practice across OTPs?** |  |
| 1. OTPs overall increased frequency of take-home doses, but only a minority of patients were granted the maximum allowable 14 or 28-day take-homes | Hunter, 2021; Krawczyk, 2022; Brothers, 2021; Figatt, 2021; Saloner, 2022; Jacka, 2021; Joseph, 2021; Amram, 2021; Amram, 2022; McIlveen, 2021; Levander, 2022 |
| 2. Take-home eligibility was most often based on patient substance use, time in treatment, ability to safely store methadone, and risk of COVID-19 | Amram, 2022; Hoffman, 2022; Krawczyk, 2022; Hunter, 2021; Brothers, 2021; Levander, 2022; Goldsamt, 2021; Kidorf, 2021; Joseph, 2021; Suen, 2022 |
| 3. Barriers to implementation of flexibilities included concerns around managing take-homes for patients with ongoing substance use, concerns around government oversight and liability, and concerns around financial sustainability | Suen, 2022; Joseph, 2021; Goldsamt, 2021; Levander, 2022; Krawczyk, 2022; Hunter, 2021; Madden, 2021 |
| 4. Facilitators to implementation of flexibilities included change agents who encouraged uptake of take-homes, leveraging multi-disciplinary clinical teams to determine take-homes, telehealth and reduced toxicology testing to support decrease frequency of visits, and using medication lock-boxes and other diversion prevention tactics | Goldsamt, 2021; Joseph, 2021; Brothers, 2021; Krawczyk, 2022; Kiforf, 2021; Dunn, 2021; Treitler, 2022 |
| **2. How did the new methadone take-home flexibilities influence perceptions and experiences of methadone patients?** |  |
| 1. Despite new flexibilities, some patients expressed continued lack of access to methadone take-homes privileges, which was often seen as unjust, burdensome and a disincentive for methadone engagement | Suen, 2022; Harris, 2022; Krawczyk, 2021; El-Bassel, 2022; Levander, 2021; Nobles, 2021; Sarker, 2022 |
| 2. Many patients expressed that increased take-homes supported their treatment experience by improving self-esteem and autonomy, reducing treatment burden, and avoiding negative triggers associated with clinic attendance | Suen, 2022; Levander, 2021; Hoffman, 2022; Harris, 2022; Walters, 2022; Nobles, 2021; Krawczyk, 2021 |
| 3. Some patients expressed that extended take-home flexibilities disrupted their routine and treatment stability, but incidences of diversion were rare | Levander, 2021; Krawczyk, 2021; Nobles, 2021; Harris, 2022; Figgatt, 2021 |
| **3. How did the new methadone take-home flexibilities influence perceptions and experiences of methadone providers?** |  |
| 1. Many providers expressed that take-home flexibilities allowed them to provide more patient-centered care, improved patient motivation, and reduced treatment burden | Suen, 2022; Hunter, 2021; Goldsamt, 2021; Treitler, 2022; Madden, 2021; Krawczyk, 2022 |
| 2. Many providers expressed concern that pandemic flexibilities and less frequent contact with patients could be destabilizing and lead to undesirable patient behaviors | Suen, 2022; Levander, 2022; Hunter, 2021; Madden, 2021; Goldsamt, 2021; Treitler, 2022; |
| **4. How did the new methadone take-home flexibilities impact overdose risk?** |  |
| 1. No significant increases in methadone overdoses were observed in relation to implementation of take-home flexibilities | Joseph, 2021; Ezie, 2022; Amram, 2021; Brothers, 2021; Welsh, 2022; Jones, 2022 |
| **5. How did the new methadone take-home flexibilities impact illicit drug use and methadone non-compliance?** |  |
| 1. Findings on changes in illicit substance use during the pandemic were mixed, but could not be necessarily attributable to new flexibilities in take-home doses | Ezie, 2022; Bart, 2022; Hoffman, 2022 |
| 2. Findings on changes in methadone non-compliance during the pandemic were mixed, but could not be necessarily attributable to new flexibilities in take-home doses | Amram, 2021; Ezie, 2022; Bart, 2022; Dunn, 2021; Kidorf, 2021 |
| **6. How did the new methadone take-home flexibilities impact methadone treatment initiation and retention?** |  |
| 1. Only one study assessed treatment retention, finding modest improvements in retention associated with increased take-home doses | Hoffman, 2022 |

### b. Findings and implications for federal regulators

#### i. Demonstrating need for the rule: uptake of take-home flexibilities

Under EO 12866, the regulator begins by explaining the need for the proposed rule.^19^ Four findings on this subject emerged: First, all studies that explored the frequency of take-home doses observed some increase following the pandemic guidance. The proportion of patients who received take-home increases varied by study and clinic. Three studies asked this question directly to OTP providers: In multi-state interviews with 20 OTP clinicians, 72% indicated that their OTPs increased the volume of take-home doses (Hunter, 2021).^21^ Similarly, 66% of OTP directors surveyed in Pennsylvania noted they extended take-home supplies following the new flexibilities (Krawczyk, 2022).^22^ In a survey of all 8 Connecticut OTPs, directors reported that the number of patients receiving one or no take-home doses decreased from 37.5% to 9.6% (Brothers, 2021).^23^

Three surveys asked patients about increased take-homes: In a survey of 104 OTP patients in North Carolina, 91.6% said they received some take-homes following the pandemic flexibilities relative to 68.3% who had received take-homes prior to the pandemic (Figgatt, 2021).^24^ In a multi-state survey, 76% of OTP patients reported receiving more take-home doses since the pandemic (Saloner, 2022).^25^ Across 8 New England OTPs, 42% of patients surveyed reported receiving increased access to take-home medication (Jacka, 2021).^26^ Other studies analyzed OTP patient records directly pre and post pandemic flexibilities: A study of five New York City OTPs saw a reduction in the proportion of patients who came to the OTP 5-6 days weekly from 47.2% to 9.4% (Joseph, 2021).^27^ A Washington OTP reported over 90% of patients experienced increases in take-home doses, with an average change of 11.4 to 22.3 monthly take-homes (Amram, 2021),^28^ with sustained increases (Amram, 2022).^29^ A study of patient records across Oregon’s 20 OTPs found a 54% reduction in mean monthly visits, with mean take-home doses increasing from 5.8 to 11.3 per month (McIlveen, 2021).^30^ This uptake suggests many providers were able and willing to implement program changes to increase take-home supply, even in a time of great uncertainty.

Second, despite some increase in take-home doses, providers did not uniformly grant patients the maximum supplies of 14 or 28 days. In a multi-state survey of 170 OTP providers, 47% reported they routinely allowed 14 days of take-home doses for newly enrolled patients, 52% allowed 14 days of take-home doses for “less stable” patients, and 66% allowed 28 days of take-home doses for “stable” patients (Levander, 2022).^31^ State-specific studies support these findings: Across Connecticut OTPs, the proportion of patients receiving 14-day take-homes increased from 14.2% to 26.8% and the proportion receiving 28-day take-homes increased from 0.1% to 16.8% (Brothers, 2021).^23^ Similarly, over 90% of OTPs in Pennsylvania noted less than half of their patients received 14-day take-homes, and 95% noted less than a quarter of their patients received 28-day take-homes (Krawczyk, 2022).^22^ No studies identified differences in take-home trends by provider characteristics (Lavender 2022, Krawczyk 2022).^22,31^ Findings suggest that even under a more permissive policy, providers will not necessarily permit all patients to receive maximum amounts of take-home supplies.

Third, some providers expressed concern over patients continuing to use other drugs, such as sedatives (Suen, 2022; Joseph, 2021).^27,32^ Some expressed concerns about reduced vigilance or oversight of patients (Goldsamt, 2021),^33^ or feared scrutiny of practices and outcomes by federal and state agencies (Levander, 2022).^31^ Many expressed concerns about how take-home dosing would reduce revenue (Krawczyk, 2022, Levander, 2022, Goldsamt, 2021, Hunter, 2021).^22,31,33,34^ Some noted concerns around legal liability for potential overdose or diversion of methadone, which made them apprehensive about take-home dosing long-term (Levander 2022, Hunter, 2021, Madden, 2021).^31,34,35^ These findings suggest provider uncertainty about the regulations, consequences for patients, and finances depressed uptake.

Fourth, despite various uncertainties, some studies described strategies to support effective uptake. This included having OTP directors act as change agents (Goldsamt, 2021),^33^ or interdisciplinary teams to guide take-home decisions (Joseph, 2021).^27^ Some studies found that providers were willing to provide increased take-homes indefinitely: In a survey of OTP directors in Pennsylvania, 79% agreed with maintaining more flexibility on take-home length (Krawczyk, 2022).^22^ Some providers believed that criteria for determining take-home doses prior to the pandemic were too strict, placing limits on providers’ clinical judgment (Treitler, 2022).^36^ Many were supportive of retaining the flexibilities to improve access to and quality of care (Suen, 2022),^32^ or worried about returning to a more restrictive schedule if the pandemic flexibilities were rescinded (Treitler, 2022).^36^ These findings support the idea that, if continued, flexibility for take-homes could become a part of regular practice.

### ii. Potential benefits of the proposed rule

The next step in regulatory analysis explores benefits and costs of the proposal. The review sheds light on the following potential benefits of long-term flexibility for take-home doses: First, many patients described that receiving increased take-homes and being given the responsibility to manage their medication resulted in feelings of pride, accomplishment and self-confidence that supported treatment goals and sobriety, and helped build a stronger relationship with their providers (Suen, 2022; Levander, 2021; Hoffman, 2022; Krawczyk, 2021).^37,32,38,39^ Patients described additional take-homes as liberating (Harris, 2021)^40^ and valued how increased take-homes, and reduced OTP visits, provided them with a sense of normalcy and stability (Levander, 2021)^38^ and reduced stigma associated with frequent clinic attendance (Walters, 2022).^41^

Patients also reported that reduced travel to the clinic gave them more time to attend to aspects of their lives such as jobs, school, caregiving, and recreation (Suen, 2022; Hoffman,2022; Walters, 2022; Nobles, 2021).^32,39,41,42^ Some described how liberating it was to not have to arrange child care or get up early and commute before or after work, to have more time for family, and spend less time and money driving to and from the clinic on a daily basis (Levander, 2021).^38^ Patients also described that increased take-homes allowed them to avoid the clinic and triggers for use: fewer clinic visits reduced exposure to individuals less stable in recovery and other potential triggers (Hoffman, 2022; Levander, 2021).^38^ Others described that having fewer people in the waiting room and reduced crowding created a healthier mental health atmosphere, and was beneficial in preventing transmission of COVID and other infections to family members (Levander, 2021).^39^

In addition to positive patient experiences, one study found that increased take-homes were associated with lower probability of treatment discontinuation: Of three groups examined by days in treatment at the start of the study period (<90, 90-180, 180+), only individuals with 90 days of treatment received increased take-homes and this group saw a significantly reduced odds of treatment discontinuation of 0.97 for every 1% increase in take-home dosing above expected pre-pandemic regimens. Only patients in treatment fewer than 90 days - which did not receive increases in take-homes - were more likely to discontinue treatment in the COVID period (13% pre-COVID vs. 26% post) (Hoffman, 2022).^39^

Beyond patient benefits, many providers expressed appreciation for the new flexibility to make better and more equitable decisions to support needs of their patients (Suen, 2022; Hunter, 2021; Goldsamt, 2021; Treitler, 2022).^32–34,36^ Some providers also noted that the increased take-home doses during the pandemic permitted greater adherence to treatment and improved autonomy and motivations to change (Treitler, 2022).^36^ Providers expressed that over-regulation of methadone undermined patient-centered care, impeded methadone access and was a waste of resources (Madden, 201; Hunter, 2021).^23^ In a survey of OTP directors in Pennsylvania, 96% agreed that take-home methadone is less burdensome for accessing treatment (Krawczyk 2022).^22^ Some OTPs were willing to try different protocols and technologies to improve flexibility, such as telehealth (Brothers, 2021)^23^ and reduced toxicology testing (Joseph, 2021)^27^ to support decreased frequency of visits.

### iii. Potential costs of the proposed rule

Some studies attempted to elucidate potential costs of flexibility for take-home doses, including concerns about overdose, patient destabilization, and diversion. Indeed, studies found that providers worried that less frequent contact with patients would lead to patient destabilization (Suen, 2022; Levander 2022)^31,32^ or difficulty building rapport (Hunter, 2021).^34^ A few expressed concerns regarding overdose risk associated with misuse (Madden, 2021; Goldsamt, 2021).^33,35^ Other concerns centered on having less control over what were seen as undesired behaviors, such as patients not taking methadone as prescribed, diverting medications, or continuing to use substances (Madden, 2021; Treitler, 2022; Goldsamt, 2021).^33,35,36^ However, these concerns rarely precipitated.

First, the review finds no significant evidence of increased methadone overdose risk as a result of the guidance. Six studies assessed the impact of the pandemic flexibilities on methadone-related overdoses: Three used OTP patient records to assess overdose events, none of which found significant increases: In a study of 3600 OTP patients, six non-fatal and no fatal overdoses were reported following the COVID flexibilities, relative to two non-fatal and one fatal overdose prior to the flexibilities (Joseph, 2021).^43^ In another study, only one of 129 OTP patients (0.7%) reported an overdose relative to three (2%) pre-pandemic (Ezie, 2022).^43^ A study of 183 patients found no significant changes in emergency department overdose visits (16 (8.7%) versus 15 (8.1%) in pre vs. post COVID-19 periods (Amram, 2021).^28^ A study assessing mortality data in Connecticut pre and post pandemic changes found that neither methadone-only nor methadone-involved fatalities increased in the five-month period in 2020 compared to earlier years, after accounting for the increase in overall fatal overdoses (Brothers. 2021).^23^

Finally, two studies analyzed data at the national level. The first analyzed data on calls to 55 poison control centers across the U.S, and found that while the number of yearly adult intentional exposures involving methadone increased by 5.3% (1,199 to 1,262), there was no significant change in reported methadone-involved hospitalizations or deaths (Welsh, 2022).^44^ The second study analyzed national data on overdose deaths, and found the proportion of methadone-involved deaths did not increase following the COVID-19 changes, despite an increase in number of overdose deaths overall during that time period (Jones, 2022).^45^ In a few qualitative studies, providers admitted that they did not observe overdoses as a consequence of increased take-homes, despite what they had anticipated (Suen, 2022; Hunter, 2021; Treitler, 2022).^32,34,36^

A second expressed concern was that take-homes would disrupt patient care routines and lead to adverse patient outcomes. The review revealed these experiences varied significantly across patients and were less commonly expressed than positive sentiments. Some patients reported feeling overwhelmed, not trusting themselves, feared the temptation to overuse their medication (Levander, 2021; Krawczyk 2021),^37,38^ or thought take-homes fractured their daily routine and sense of stability (Harris, 2021).^42^ Some expressed difficulty adhering to the prescribed dosing regimen and feared that admitting this to the OTP would result in losing their new take-home privileges (Nobles, 2021).^42^

Other studies assessed patient stability by looking at patient non-compliance with methadone, as established by urine toxicology. One study found no significant change in the number of OTP patients with a negative methadone screen (15.8% vs. 16.9%) (Amram, 2021),^28,42^ and another found no significant change in positive tests for methadone (92% vs 96%) in the pre and post-pandemic periods (Ezie, 2022).^43^ Only one OTP study found a significant increase in the percent of tests negative for methadone (1.9% vs. 4%) (Bart, 2022).^46^ Despite these mixed findings, across all studies, methadone non-compliance remained rare even after implementing take-home flexibilities.

A final set of studies considered destabilization by assessing changes in illicit substance use. The findings were also mixed with one finding no statistically significant change between pre and post-periods in the positive test detection for non-prescribed opiates (39 to 36%) or other illicit drugs (45% to 40%) (Ezie, 2022),^43^ while another finding an increase in positive tests for opiates (14 to 22%), benzodiazepines (6.3% to 11%), and methamphetamine (10% to 16%) in (Bart, 2022).^46^ A study analyzing changes in drug use by patient time in treatment found only patients in treatment between 90-180 days, who did not receive an increase in take-homes during the study period, saw an increase in other drug use from 19 to 33% (Hoffman, 2022).^39^ This implies increases in illicit substance use are not necessarily attributable to increased take-homes.

Relatedly, some studies explored whether increased take-home doses increase diversion. “Diversion” is not defined in federal law but generally means the “selling/trading, sharing or giving away,” either voluntarily or involuntarily (e.g., by way of theft), of a prescription medication to someone to whom it was not prescribed.^47^ Only one reviewed study surveyed OTP patients directly about diversion of methadone take-homes. Only 14.4% (n=15) reported knowing someone who gave away doses, most commonly noted to be as a result of needing money or drugs (38.5%), helping someone else (37.5%), or saving up for travel (28.8%) (Figatt, 2021).^24^ In some cases, providers admitted that their concerns about diversion did not materialize (Treitler, 2022; Brothers, 2021).^23,36^ Many studies also described tactics OTPs implemented to reduce diversion, such as medication lock-boxes (Krawczyk 2022, Dunn, 2021; Kidorf, 2021),^22,48,49^ and medication callbacks (Tetiler, 2022; Krawczyk 2022).^22,36^

### iv. Alternatives and their implications

Next, the regulator must work through alternative formulations of a proposed change and consider their implications.

One core issue for SAMHSA’s rule will be defining which patients may receive additional take-home supplies. One option would be that SAMHSA declines to restrict the amount take-home supplies.^50^ This approach would default to providers adopting a medical standard of care rather than proscriptive federal rules. The review suggests that dosing decision freedoms would be treated with caution by providers; as most OTPs declined to provide maximum take-home supplies to patients in the context of the pandemic. Another option would be for SAMHSA to propose an undefined standard like “stable” or “less stable” that stops short of deferring to the medical standard of care but allows providers to exercise subjective judgment. This would mirror the first 20 months after SAMHSA initially provided take-home flexibilities and before it issued guidance with more specific criteria. A final option would be for SAMHSA to provide detailed definitions of “stable” and “less stable” in regulation. In November 2021, SAMHSA issued additional guidance that provided more explicit criteria, including the requirement for 60 days of negative toxicological screening. SAMHSA might therefore be expected to propose these additional criteria to continue the pandemic policy.

A second issue is whether patients have recourse to appeal take-home decisions. When regulators craft policies that will result in some patients receiving more flexible treatment options than others, based on subjective criteria (e.g., “stable” or “less stable”), patients might reasonably have concerns about whether they are being treated fairly. The review reveals that, in the context of pandemic flexibilities, patients often viewed their lack of access to take-homes as unjust, discriminatory, burdensome, and expressed concern at being required to come to the clinic daily even in the midst of a pandemic (Krawczyk 2021, Harris, 2021; El-Bassel, 2022).^37,40,51^ Some patients with ongoing substance use or who lacked housing were frustrated that they were excluded from take-home privileges, and others voiced frustrations about being given increased take-home doses that were subsequently rolled back (Lavender 2021; Suen, 2022; Harris, 2021).^32,38,40^ Some were denied additional doses and not believed by staff when their doses had spilled, lost, or were stolen from them (Nobles, 2021; Harris, 2021).^40,42^ The review shows that, as a result of such frustrations, some individuals felt they had to self-manage withdrawal or reported that they wished to stop methadone treatment altogether (El-Bassel, 2022; Krawczyk 2021).^37,51^

These experiences described align with long-running complaints about care provided by OTPs among methadone patients.^14^ In designing its new take-home rules, SAMHSA could help address these issues, such as allowing patients to appeal an OTP’s decision around take-homes or otherwise request a second opinion. Because of the scarcity of OTPs and how tightly methadone is regulated, patients do not always have realistic options for alternative providers. While adding more methadone provider and treatment setting (e.g., office-based methadone prescribing) options may ultimately be the ideal remedy, an oversight process for patient care decisions is an alternative that could mitigate problematic provider behavior, where appropriate, and therefore give patients confidence that they are being treated fairly.

## 4. Discussion

The review provides key evidence for SAMHSA to consider as it takes steps to make permanent the methadone flexibilities it made available in response to the COVID-19 pandemic. Importantly, findings from research evidence suggest this change did not result in significant overdoses or other adverse effects among patients. On the contrary, data indicates potentially improved treatment retention, substantial quality of life and self-efficacy improvements, reduced burden, and fewer stressful clinic encounters for patients were associated with greater take-home flexibility. Benefits were also described among treatment providers, including improved patient motivation and satisfaction in the ability to provide patient-centered care. Therefore, efforts to create a permanent policy to support increased flexibility of take-home doses, with the goal of improved patient outcomes, is well-supported.

The review finds that, once offered, many providers took up this new flexibility, suggesting that they will use ongoing flexibility to benefit patients. Providers did not, however, default to providing maximum take-home supplies to patients, which should allay some implementation concerns. Uncertainty tended to depress uptake, which suggests the importance of a permanent change, as provider attitudes in light of greater certainty will likely continue to contribute to uptake. In proposing new rules to extend the benefits of greater take-home flexibilities more permanently, SAMHSA has two main choices: Whether to dictate which patients qualify for additional flexibility and how this flexibility should be determined, and whether patients have recourse to appeal take-home decisions. The review sheds light on those choices.

The review also sheds light on key implementation issues. First, the review suggests that SAMHSA would be well-advised to expect and plan for provider uncertainty. While many providers acknowledged the benefits of increased take-home doses and the flexibility it allotted them, many also expressed hesitancy about how the new take-home allowances would work. In a complex area of patient care subject to a wide range of different legal requirements, if SAMHSA’s goal is to encourage uptake of additional flexibilities, providers might require technical assistance and implementation support to work through their concerns without fear of penalty.

A second implementation factor to consider is the role of states in choosing to embrace the pandemic flexibilities, as some states declined to take advantage of these flexibilities,^52^ partially attributed to their temporary nature.^53^ A long-term regulatory change could therefore make it more likely that additional states/jurisdictions would take up the flexibility. Other states have objected to the flexibility on policy grounds, preferring to keep the status quo approach to OUD treatment with methadone.^53,54^ As federal regulations give states a large role in overseeing OTPs, SAMHSA could consider how to proactively support and encourage implementation of this policy at the state level,^13^ and how to oversee and ensure individuals providers are complying with such regulations. Issuing stable policy rather than iterative guidance could reduce uncertainty. Providing technical assistance to address provider questions is another. A third approach could be to increase SAMHSA’s oversight of the relevant state regulators that oversee OTPs.

Another third consideration is the potential utility of diversion-prevention tactics such as use of lock boxes. These approaches are not without their problems. For example, lockboxes may not be feasible for homeless populations and over-reliance on urine drug tests can be burdensome and sacrifice patient-provider trust.^14^ Additional study, including pilot testing, could help SAMHSA make informed decisions about whether these emerging approaches strike the right balance between equitable care and concerns about safety and diversion.

Lastly, while payment policy issues are likely outside of SAMHSA’s discretion, providers demonstrated awareness of and sensitivity to the financial implications of changes to service delivery models. To the extent that SAMHSA policymakers can factor this into their consideration of alternatives, including making recommendations to other agencies to align their policies and payment systems, it may help ensure that implementation aligns with policy goals.

Turning to the limitations of our review and analysis, our search was confined to peer-reviewed studies published before September 2022, and may have missed more recent studies or studies published in the gray literature. Furthermore, synthesis and interpretation of evidence was limited by the wide variation in settings, outcomes, and methods used to answer research questions of interest. For instance, observational studies that used clinical data from OTPs varied greatly in size, the methods and types of data used to assess outcomes, and time periods assessed in relation to the implementation of the new COVID-19 regulations. Moreover, clinics that conducted their own evaluations or agreed to participate in research likely represent academic or research-oriented programs, and their practices, providers and patients may not represent OTPs more broadly. Findings should also be interpreted while considering possible social desirability bias involved in self-report surveys and interviews, and potential confounders not accounted for in quantitative data studies. Importantly, the quickly changing nature of the pandemic, and the multiple associated social and structural changes make it difficult to attribute outcomes directly to SAMHSA’s take-home guidance. This includes the potential influence of other changes to OTP practices such as use of virtual platforms for behavioral services and less urine drug screening, which were not explored in this review. As such, none of the associations described above can be determined to be causal.

Despite these limitations, our review proposes concrete policy considerations based on evidence triangulated from across diverse research settings, geographies, populations, data sources and stakeholder groups. Particularly important is the integration of the perspectives of methadone patients as gathered by qualitative research, as patients at the center of the substance use treatment system are often excluded from these important policy conversations. Many questions remain around the impact of this policy change, including the role of other methadone delivery practices such as counseling and drug screening on the experiences and health of methadone patients, and how technologies such as safety boxes and virtual health platforms can aid new regulatory environments. There are also many larger discussions to be explored around federal versus state role for regulators, and the role of OTPs in the delivery of methadone more broadly: This includes the potential of expanding methadone treatment to an office-based or pharmacy-dispensing delivery system as is done in other countries,^55^ and how that would impact access and the experience of patients.^56^ These questions should be the subject of further research and ongoing discussion but should not act as a deterrent to timely implementation of changes for which there is strong evidence so far, including the many observed benefits and few drawbacks of greater take-home flexibilities, gathered by existing studies.

## Conclusion

It took a pandemic to break through long-standing rules that have constrained patient access to methadone in the U.S. Returning to the pre-pandemic status quo would forgo the considerable benefits discussed above. While any policy that makes it incrementally easier for patients to self-administer methadone opens the door to certain risks, those risks trade off against significant benefits. On balance, based on the pandemic experience captured in this review, a more flexible approach to take-home medication will be net beneficial for patients and society as a whole, and is urgently needed during this ongoing overdose crisis.

## Data Availability

All data produced in the present study are available upon reasonable request to the authors

**Appendix Table 1.** Search strategy by database.

| Database | Strategy |
| --- | --- |
| **PubMed** | 1 ("Methadone" AND ("take-home" OR "take-home")) |
|  | 2 1 OR "Methadone maintenance therapy" OR "OTP" OR "Opioid treatment program" |
|  | 3 2 OR "Opiate Substitution Treatment"[MeSH Terms] |
|  | 4 "Methadone" OR "Methadone maintenance therapy" OR "OTP" OR "Opioid treatment program" OR "Opiate Substitution Treatment"[MeSH Terms] |
|  | 5 "Methadone" OR "Methadone maintenance therapy" OR "OTP" OR "Opioid treatment program" |
|  | 6 ("COVID-19"[Mesh]) AND ("Methadone" OR "MMT" OR "Methadone maintenance therapy" OR "methadone maintenance treatment" OR "OTP" OR "Opioid treatment program") |
|  | 7 6 OR "take-home" OR "take-home" |
|  | 8 ("COVID-19"[Mesh] OR "Pandemics"[Mesh]) AND ("Methadone/therapeutic use"[Mesh] OR "Methadone/administration and dosage"[MeSH] OR "MMT" OR "Methadone Maintenance Therapy" OR "Methadone Maintenance Treatment" OR "OTP" OR "OTPs" OR "Opioid treatment program" OR "opioid treatment programs" OR "take-home" OR "Take-home" OR "Opioid-Related Disorders/drug therapy"[Mesh] OR "Opioid-Related Disorders/therapy"[Mesh] OR "Opiate Substitution Treatment"[MeSH Terms] OR "Opioid-Related Disorders/prevention and control"[Mesh]) |
|  | 9 8 AND 1 |
| **PsycInfo** | 1 covid-19.mp. or exp COVID-19/ |
|  | 2 exp Pandemics/ or exp Severe Acute Respiratory Syndrome/ or sars-cov-2.mp. |
|  | 3 1 or 2 |
|  | 4 exp Methadone Maintenance/ or exp Methadone/ or methadone.mp. |
|  | 5 3 AND 4 |
| **Google Scholar** | allintitle: methadone AND (pandemic OR change OR covid OR policy OR "expanded" OR "take-home" OR otp OR "opioid treatment program" OR mmt OR "maintenance therapy" OR "maintenance treatment") |

**Appendix Table 2:** Summary of reviewed articles.

|  | **First Author, Year** | **Research Questions Addressed** | **Study Design** | **Study Period (pre, post pandemic)** | **Setting** | **Study Population** | **Sample Size** |
| --- | --- | --- | --- | --- | --- | --- | --- |
| 1 | Amram, 2021 | Implementation, Overdose, Illicit drug use/diversion | Observational outcomes study | Pre: 270 days prior to Mar. 1, 2020<br>Post: 270 days after April 1, 2020 | One OTP, Spokane WA | English-speaking clients age 18 years and older receiving methadone for OUD | 183 |
| 2 | Amram, 2022 | Implementation | Observational outcomes study | Pre: Dec. 1 2019 – Feb. 29, 2020<br>Post: Apr. 1 2020 – Jun, 30 2020 | One OTP, Spokane WA | English-speaking clients age 18 years and older receiving methadone for OUD | 194 |
| 3 | Bart, 2022 | Illicit drug use/diversion | Observational outcomes study | Pre: Jul. 2019<br>Post: Jul. 2020 | One OTP, Minneapolis, MN | Clients receiving methadone for OUD | 613 |
| 4 | Brothers, 2021 | Implementation, Overdose, Illicit drug use/diversion | Observational outcomes study | OTP survey: Jul 8- Aug. 18, 2020<br>Death data: 2020<br>Pre: 2015-201 Post: Jan.- Aug. | Multiple OTPs, Mortality data, CT | Clients receiving methadone for OUD | 24,261 |
|  | Dunn, 2021 | Implementation, Illicit drug use/diversion, Patient Experience | Randomized Trial | Mar. 2018 – Mar. 2020 | One OTP, Baltimore, MD | Clients age 18 years and older receiving methadone for OUD | 25 |
| 6 | El-Bassel, 2022 | Patient experience | Quantitative content analysis | Mar. 1- May 22, 2020 | Reddit, subreddits: r/opiates, r/OpiatesRecovery, r/suboxone, and r/Methadone | Clients receiving methadone for OUD | 9,809 |
| 7 | Ezie, 2022 | Overdose, Illicit drug use/diversion | Observational outcomes study | Pre: Dec. 16, 2019 – Mar. 15, 2020<br>Post: Mar. 16- June 15, 2020 | One OTP, New York, NY | Veterans receiving methadone for OUD | 129 |
| 8 | Figgatt, 2021 | Implementation, Illicit drug use/diversion | Closed end survey of practices/perceptions | Pre/Post March 1, 2020 | Multiple OTPs, Greensboro, NC | Clients receiving methadone for OUD | 104 |
| 9 | Goldsamt, 2021 | Implementation, Provider experiences | Qualitative study | "Early pandemic" | Multiple OTPs, Multi-state | MOUD Providers | 25 |
| 10 | Harris, 2022 | Patient experience | Qualitative study | Aug. – Oct. 2020 | One OTP, Boston, MA | Adults 18-65 with overdose in last 3 years | 20 |
| 11 | Hoffman, 2022 | Implementation, Illicit drug use/diversion, Initiation/Retention Tx, Patient Experience | Observational outcomes study ; qualitative interviews | Pre: Sep. 1, 2019 – Feb. 28, 2020<br>Post: Apr. 1, 2020 – Sep. 30, 2020 | Multiple OTPs, OR | Clients receiving methadone | 377 (quant)<br>32 (qual) |

|  | **First Author,**<br>**Year** | **Research Questions**<br>**Addressed** | **Study Design** | **Study Period**<br>**(pre, post pandemic)** | **Setting** | **Study Population** | **Sample**<br>**Size** |
| --- | --- | --- | --- | --- | --- | --- | --- |
| 12 | Hunter, 2021 | Implementation,<br>Provider experiences | Qualitative study | May 14, 2020 - June 19, 2020 | Multiple OTPs,<br>Multi-state | MOUD Providers | 20 |
| 13 | Jacka, 2021 | Implementation | Observational<br>outcomes study | May - July 2020 | Multiple OTPs,<br>Multi-state | Clients age 18 years +<br>receiving methadone for OUD | 135 |
| 14 | Jones, 2022 | Overdose | Observational<br>outcomes study | Pre: Jan. 2019- Feb 2020<br>Post: Apr. 2020- Aug 2021 | Mortality data,<br>Multi-state | Adults 18 + | N/A |
| 15 | Joseph, 2021 | Implementation,<br>Overdose | Observational<br>outcomes study | Mar. 16, 2020 - May 31, 2020 | Multiple OTPs,<br>Bronx NY | Clients receiving methadone | 3,600+ |
| 16 | Kidorf, 2021 | Implementation, Illicit<br>drug use/diversion | Observational<br>outcomes study | Apr. 8– Jul. 6, 2020 | One OTP,<br>Baltimore, MD | Clients receiving methadone | 42 |
| 17 | Krawczyk, 2021 | Patient experience | Qualitative study | Mar. 5- May 13, 2020 | Reddit: subreddits:<br>r/opiates,<br>r/OpiatesRecovery, | Clients receiving methadone | 2,000 |
| 18 | Krawczyk, 2022 | Implementation,<br>Provider experiences | Closed end survey | Sep.- Nov. 2020 | Multiple OTPs,<br>PA | Clinical directors of OTPs | 47 |
| 19 | Levander, 2021 | Patient experience | Qualitative study | Pre: Aug.- Oct. 2020<br>Peri: Nov. 2020 – Jan. 2021 | Multiple OTPs,<br>OR | English-speaking clients age<br>18+ receiving methadone | 46 |
| 20 | Levander, 2022 | Implementation,<br>Provider experiences | Closed end survey | Sep. – Nov. 2020 | Multiple OTPs,<br>Multi-state | OTP leaders | 170 |
| 21 | Madden, 2021 | Provider experience | Qualitative study | Feb. 17, 2017 – Aug. 31, 2020 | Multiple OTPs,<br>Multi-state | OTP staff | 59 |
| 22 | McIlveen, 2021 | Implementation | Observational<br>outcomes study | Pre: Feb. and first half of Mar, 2020<br>Post: Mar-Jun 2020 | Multiple OTPs,<br>OR | Clients receiving methadone | 7,792 |
| 23 | Nobles, 2021 | Patient experience | Qualitative study | Jan. 31- Sep. 30, 2020 | Reddit Subreddits: r/<br>methadone | Clients receiving methadone | 215 |
| 24 | Saloner, 2022 | Implementation | Closed end survey | Aug.19, 2020 – Jan. 29, 2021 | Convenience sample of<br>substance use treatment and<br>harm reduction programs,<br>Multi-state | Individuals receiving substance<br>use treatment | 243 |
| 25 | Sarker, 2022 | Patient experience | Quantitative content<br>analysis | Pre: Jan. 1, 2019 – Feb. 29, 2020<br>Peri: Mar. 1, 2020 – Nov. 30, 2020 | Reddit | Clients receiving methadone | 820 |
| 26 | Suen, 2022 | Implementation,<br>Provider experiences,<br>Patient experience | Qualitative study | Provider: Aug, to Sep, 2020<br>Patient: Sep. to Nov. 2020 | One OTP,<br>San Francisco, CA | MOUD providers, clients<br>receiving methadone | 10<br>providers,<br>20<br>patients |
| 27 | Treitler, 2022 | Provider experience | Qualitative study | Sep. – Nov. 2020 | Providers from OTPs or<br>office-based treatment, NJ | MOUD providers | 20 |
| 28 | Walters, 2022 | Patient experience | Qualitative study | Jun. – Oct.2020 | Substance use<br>clinics/programs and<br>craigslist ads, Northeast | Adults 18+ who use/used<br>drugs; MOUD providers, clinic<br>staff, or work at a regulatory<br>agency | 21 clients<br>18<br>providers,<br>or gov.<br>agencies |
| 29 | Welsh, 2022 | Overdose | Observational<br>outcomes study | Pre: Mar. 19, 2019 – Mar. 16, 2020<br>Post: Mar. 17, 2020 – Mar. 15, 2021 | Poison Control Center Data,<br>Multi-state | Adults ≥ 18 years old | 2,461 |

